# Prediction of Outcomes in Infants with Hydrops Fetalis by Mode of Delivery: A Retrospective Cohort from a Low-Resource Setting in Kenya

**DOI:** 10.1101/2024.11.08.24316964

**Authors:** Philippe P. Amubuomombe, Wycliffe K. Kosgei, Philiph Tonui K., Richard M. Mogeni, K. Mutindi, Sarah K. Esendi, Ruth Ngeleche, Paul Nyongesa, Irene Koech, Jignesh K. Jesani, Esther Wanjama, Rajshree K. Hirani, Emily Chesire, Donah Oeri, Audrey K. Chepkemboi, Deborah V. Makasi, Vahista J. Shroff, Bett C. Kipchumba, Pallavi Mishra, Philip Kirwa, Amgad Hamza, Wilson K. Aruasa, Ann Mwangi, Elkanah O. Orang’o

## Abstract

**BACKGROUND:** Hydrops fetalis is a condition associated with increased perinatal and neonatal mortality and morbidity. The overall survival rate of infants diagnosed with hydrops fetalis is currently estimated to be 27%, despite advanced intrauterine and neonatal care. Factors that contribute to poor perinatal and neonatal outcomes have been identified; however, little is known about the existing specific tool for predicting perinatal outcomes by mode of delivery.

**OBJECTIVE:** This study aimed to determine whether cesarean section improves the perinatal outcomes of infants with hydrops fetalis in low-resource settings.

**STUDY DESIGN:** This was a retrospective cohort study in which 102 medical records of pregnancies complicated by hydrops fetalis were retrieved. For all included women, transabdominal ultrasound was performed during pregnancy as part of the standard diagnostic modality for hydrops fetalis. The medical records of all pregnant women and their newborns were retrieved and reviewed to collect information related to the outcomes by mode of delivery, either cesarean section or vaginal delivery. The pregnant women were divided into the following 4 classes based on the severity of hydrops fetalis determined by obstetric ultrasound findings: class I (mild features of hydrops fetalis), II (moderate features of hydrops fetalis), III (moderately severe features of hydrops fetalis), and IV (severe features of hydrops fetalis). The significance of the obtained data was set at a two-tailed p<0.05.

**RESULTS:** This cohort study estimated the proportion of hydrops fetalis to be 0.8%. Nonimmune hydrops fetalis was the common type, accounting for 75.6% of all cases. The 7-day survival rate was estimated to be 42.2%. There was no statistically significant association between the mode of delivery and perinatal survival (pv=0.84). Survival increased, especially in class II patients (47.5%), while survival was similarly low between class III and IV patients (22.5%). A statistically significant association was between class and survival (pv <0.001).

**CONCLUSION:** Cesarean section delivery does not improve the perinatal outcomes of hydrops fetalis infants. Creating a validated tool for predicting the perinatal outcomes of infants with hydrops fetalis by mode of delivery is useful for assisting in decision-making and predicting perinatal outcomes.

## Introduction

Hydrops fetalis is a serious life-threatening syndrome characterized by an abnormal accumulation of fluid in two or more fetal compartments, including ascites, pleural effusion, pericardial effusion, and skin edema^1,4^. Studies have shown that nonimmune causes account for up to 90% of all cases^5,8^, despite poor perinatal outcomes and low one-year survival rates ^5,9^.

The prenatal diagnosis of hydrops fetalis is difficult, especially in resource-limited settings. The overall incidence of hydrops fetalis is between 1 in 1500 and 1 in 3000 pregnancies^10,11^, and hydrops fetalis is associated with a high risk of perinatal mortality^12^. In recent publications from developed countries, the overall survival rate of infants diagnosed with hydrops fetalis was 27%^4,13^. Evidence has shown that the underlying cause of the syndrome is a predictor of poor perinatal outcomes^10^. For example, in infants with immune hydrops fetalis (IHF), a perinatal mortality rate of 3% has been reported^10^. Fifty percent of infants diagnosed in utero die, while the remaining 50% of live-born infants do not survive the neonatal period^10^. Nonimmune hydrops fetalis (NIHF) is commonly associated with severe heart failure^10^. Furthermore, evidence has shown that gestational age at birth, initial diagnosis time, birthweight, the Apgar score, and maternal comorbidities affect the perinatal survival rate^9,11^.

Globally, efforts are continuously being made to improve the perinatal outcomes of infants with hydrops fetalis through early antenatal diagnosis and in-utero therapy. Expertise in intrauterine therapeutic and genetic interventions is widely lacking in low-resource settings. Intending to improve perinatal outcomes, cesarean section has been widely performed^9^. A study performed by Unal E.T. et al.^11^, in which where 90.5% of infants with hydrops fetalis were born by cesarean section, provided evidence that pregnant women whose fetuses are diagnosed with hydrops fetalis are likely to undergo cesarean delivery despite poor perinatal outcomes and a 20-30% one-year survival rate^6,9^.

The beneficence-based obligations to fetuses in terms of classifications of fetal anomalies based on the probability of antenatal diagnosis and the probability of perinatal outcomes are fundamental to assisting obstetricians in decision-making regarding the mode of delivery of infants with hydrops fetalis^14,16^. Although treatment options for hydrops fetalis are limited, studies have shown that intrauterine fetal intervention, mainly intrauterine fetal transfusion, maternal antiarrhythmic medications for fetal arrhythmia, and in-utero surgery, such as fetal thoracentesis/paracentesis and surgical resection, are effective at improving overall survival^9,17,19^. There are several studies on hydrops fetalis given that this topic is an interesting area of continuous investigation. In this particular study, we hypothesized that cesarean delivery would improve the perinatal outcomes of pregnancies complicated by hydrops fetalis. However, there is a paucity of data on whether the mode of delivery of infants with hydrops fetalis influences perinatal outcomes, especially in low-resource settings. To fill this knowledge gap, we conducted a retrospective cohort study using the medical records of pregnant women who did and did not undergo cesarean section and their newborns 1) to determine the proportion of hydrops fetalis in a low-resource setting from the age of viability accepted in low and middle-income countries (LMIC); 2) to describe the characteristics of women whose pregnancies were complicated by hydrops fetalis; 3) to describe the clinical characteristics of newborns affected by hydrops fetalis and compare their outcomes using the developed predictor index (Predictor of Hydrops Fetalis Infant Outcomes by Mode of Delivery (PHIOMoD) Index); 4) to describe predictors of adverse perinatal outcomes in pregnancies complicated by hydrops fetalis; and 5) to determine whether cesarean delivery improves the perinatal outcomes of infants with hydrops fetalis.

## Materials and Methods

In this retrospective cohort study, the medical records of pregnant women and their newborns admitted and managed at Rely Mother and Baby Hospital of Moi Teaching and Referral Hospital (MTRH) in Kenya between 2013 and 2023 who met the following criteria were retrieved and reviewed: all pregnant women with a gestational age ≥ 28 weeks (considered age of viability in developing countries) whose fetuses were diagnosed with hydrops fetalis and whose decision for delivery was made when the fetus was alive or born alive; and all newborns treated for hydrops fetalis. Newborns with a diagnosis of in-utero death and newborns admitted to the newborn unit (NBU) for hydrops fetalis complications with missing maternal medical records were excluded from the study. To date, there is no local protocol guiding the management of pregnant women with hydrops fetalis that takes into consideration limited resources to optimize neonatal outcomes. The care for affected pregnant women, especially the mode and time of delivery is not standardized. This retrospective study attempts to address the observed gaps with a perspective to conduct a large prospective.

### Diagnostic criteria

Hydrops fetalis is mostly an incidental finding during routine prenatal workups, especially obstetric ultrasonography. In this study, the diagnostic criteria for hydrops fetalis were the identification of excess serous fluid in at least one compartment (ascites, pleural effusion, or pericardial effusion) and skin edema (thickness greater than 5 mm). Alternatively, the diagnosis was made by identifying excess serous fluid in two potential compartments without skin edema^20^. Placental edema (placental thickness > 4 cm in the second trimester and >6 cm in the third trimester) was added to this criterion^20^. In addition, the prenatal diagnosis of fetal anemia among hydrops fetalis was enhanced with a middle cerebral artery (MCA) Doppler study, which is considered a modern and accurate noninvasive diagnostic procedure. The postnatal fetal evaluation was also used for infants born with anasarca^21^ or skin edema without ultrasound findings^20^.

The developed clinical predictor tool quantified the contributions of relevant patient characteristics based on prenatal ultrasonographic findings. A focused review of the literature was conducted to identify criteria based on which the PHIOMoD study classification was developed. The strengths of the process were the creation of a multidisciplinary team (consisting of 2 maternal-fetal medicine (MFM) specialists, 8 general obstetrician-gynecologists, 2 midwives, 1 neonatologist, 1 pediatric cardiologist, 1 neonatology nurse specialist, 2 general pediatricians, and 6 senior medical officers) and the use of scientific evidence^20,22,27^ to create a comprehensive classification of hydrops fetalis based on severity and to predict the outcomes by mode of delivery based on the classification. To assess whether the PHIOMoD tool was accurate, the medical records of a few patients were selected and tested. Internal validation of the PHIOMoD tool was performed.

The standard of evidence for the optimal management of pregnant women whose fetuses are diagnosed with hydrops fetalis and management outcomes were also included. We acknowledge the variability in practice between various hospitals worldwide, experience, differences in resources, and access to intrauterine management and neonatal care. Figure 1 describes each PHIOMoD class based on the ultrasonography findings.

This ultrasound-based classification used to predict perinatal outcomes for hydrops fetalis was named the PHIOMoD Index. Maternal rhesus screening was performed as a standard to rule out maternal-fetal blood incompatibility. Maternal-fetal rhesus incompatibility (a rhesus-negative mother and rhesus-positive fetus) guided the diagnosis of IHF. All the rhesus-negative mothers underwent the indirect Coombs test (ICT) between 28 and 32 gestational weeks, and all the infants born to rhesus-negative mothers underwent the direct Coombs test (DCT). The absence of maternal-fetal rhesus incompatibility guided the diagnosis of NIHF with a negative ICT result. The etiologic diagnosis of idiopathic hydrops fetalis was made in the absence of an obvious cause of the disease.

Chromosomal testing is not performed due to limited resources available and financial constraints to the patients; however, all infants with hydrops born dead or alive were clinically evaluated, and in a few cases, investigations (postmortem and histopathology) were performed to rule out the presence of obvious congenital anomalies, such as diaphragmatic hernia, midgut volvulus, and gastrointestinal obstruction, including jejunal atresia, malrotation of the intestines, and the presence of an intraabdominal mass. Other fetal anomalies included fetal tumors, twin-to-twin transfusion syndrome, twin anemia polycythemia sequence (TAPS), congenital hydrothorax, cardiovascular anomalies such as arrhythmias, cardiomyopathy, cardiac tumors, vascular abnormalities, and placental and cord abnormalities. In addition, no intrauterine intervention or transfusion was performed in all participants because the resources were not available during the study period.

**Maternal characteristics** of interest included both sociodemographic and clinical characteristics, such as age, marital status, education level, health insurance status, parity, previous scars, history of pregnancy loss, history of isoimmune blood group antibodies, ABO and rhesus status, anemia during pregnancy, prior administration of blood products, history of illicit drug use, history of teratogenic drug use, cardiovascular disease status, thyroid disease status, diabetes mellitus status, hemoglobinopathy (sickle cell disease) status, coagulopathy status, previous fetal death, history of twin pregnancy, history of congenital malformation in a previous child, history of polyhydramnios, history of hydrops fetalis in a previous child, history of congenital heart disease in a previous child, history of feto-maternal transfusion, history of prolonged or excessive jaundice, prior neonatal death, mirror syndrome (maternal hypertension, edema, and proteinuria associated with hydrops fetalis), antibody screening results, ICT results, Kleihauer-Betke test results were done for some patients; however, serology tests, including toxoplasmosis, rubella, cytomegalovirus, herpes simplex virus, parvovirus B19 were not done due resources limited. However, the syphilis test was done for almost all patients during routine antenatal care visits. There was evidence of treatment of syphilis for some patients. Maternal characteristics were considered confounders.

**Perinatal characteristics** of interest included the following: *antenatal characteristics*: gestational age; abnormal fluid collection in ≥2 fetal compartments (ascites, pleural effusion, and pericardial effusion); skin edema ≥ 5 mm; abnormal amniotic fluid index or a maximum vertical pocket; abnormal Doppler velocimetry results (umbilical artery, middle cerebral artery, ductus venosus). Amniocentesis (for serological tests or karyotyping); cordocentesis results (for karyotyping or fetal hemoglobin or serological tests); intrauterine therapeutic interventions were not done. Placental thickness; and *postnatal characteristics*: Apgar score ≤ 5 at 5 minutes, abnormal gross morphology characteristics, abnormal placenta or placentomegaly, and abnormal echocardiography results were recorded; however, serological tests (toxoplasmosis, rubella, cytomegalovirus, herpes simplex virus, parvovirus were not done. The serology for Syphilis was done and the result was recorded. The autopsy (fetal or neonatal death), and neonatal screening results (inborn error of metabolism) were not done.

**The mode of delivery** was an independent variable. Participants’ medical records were divided into 2 arms according to the mode of delivery: cesarean delivery vs. vaginal delivery. Cesarean section was defined as an exposure. In the case of a cesarean section, indications of the cause of the cesarean section, such as hydrops fetalis, nonreassuring fetal status (defined by fetal bradycardia or tachycardia or abnormal Doppler velocimetry results), previous scars, congenital abnormalities, failed induction, cephalopelvic disproportion, ante- or intrapartum hemorrhage, and labor dystocia, were identified.

**The perinatal outcomes** of interest included low birth weight, prematurity, the Apgar score at 5 minutes, admission to the NICU, neonatal anemia status, the use of neonatal interventions such as transfusion or intubation, early neonatal death, time of death after delivery, hospital duration, cause of death, and survival status 7 days after delivery. Figure summarizes the participants enrollment process.

This study was approved by the Moi Teaching and Referral Hospital-Moi University Institutional Research and Ethics Committee (IREC) under the number FAN 0003998 (for the period from 18 October 2021 to 17 October 2022) for data collection from patient medical records and dissemination of the findings. This approval was extended up to October 2023.

### Statistical analysis

The data were entered into a computer and analyzed using the IBM SPSS software package version 20.0 (Armonk, NY: IBM Corp). Qualitative data are presented as numbers and percentages. The Shapiro‒Wilk test was used to verify the normality of the distribution.

Quantitative data are described as the range (minimum and maximum), mean, standard deviation, median, and interquartile range (IQR).

The significance of the obtained results was judged at the 5% level. Odds ratios (ORs) with 95% confidence intervals (CIs) were calculated. Binary logistic regression analyses were used to adjust the results and ORs for potential confounders. All demographic and clinical characteristics were different between the groups according to univariate analyses. Differences were considered statistically significant at P<0.05. All P values were obtained using two-tailed tests.

## Results

During the study period, there were 121,203 deliveries. During the 10 years, 102 pregnancies diagnosed with hydrops fetalis at a gestational age of 28 weeks and above were identified. The proportion of hydrops fetalis was estimated to be 0.08%. The mean maternal age was 26.8 ±6.3 (standard deviation) years, with an IQR between 22.0 and 30.0 years, as shown in Table 1.

**Table 1.**
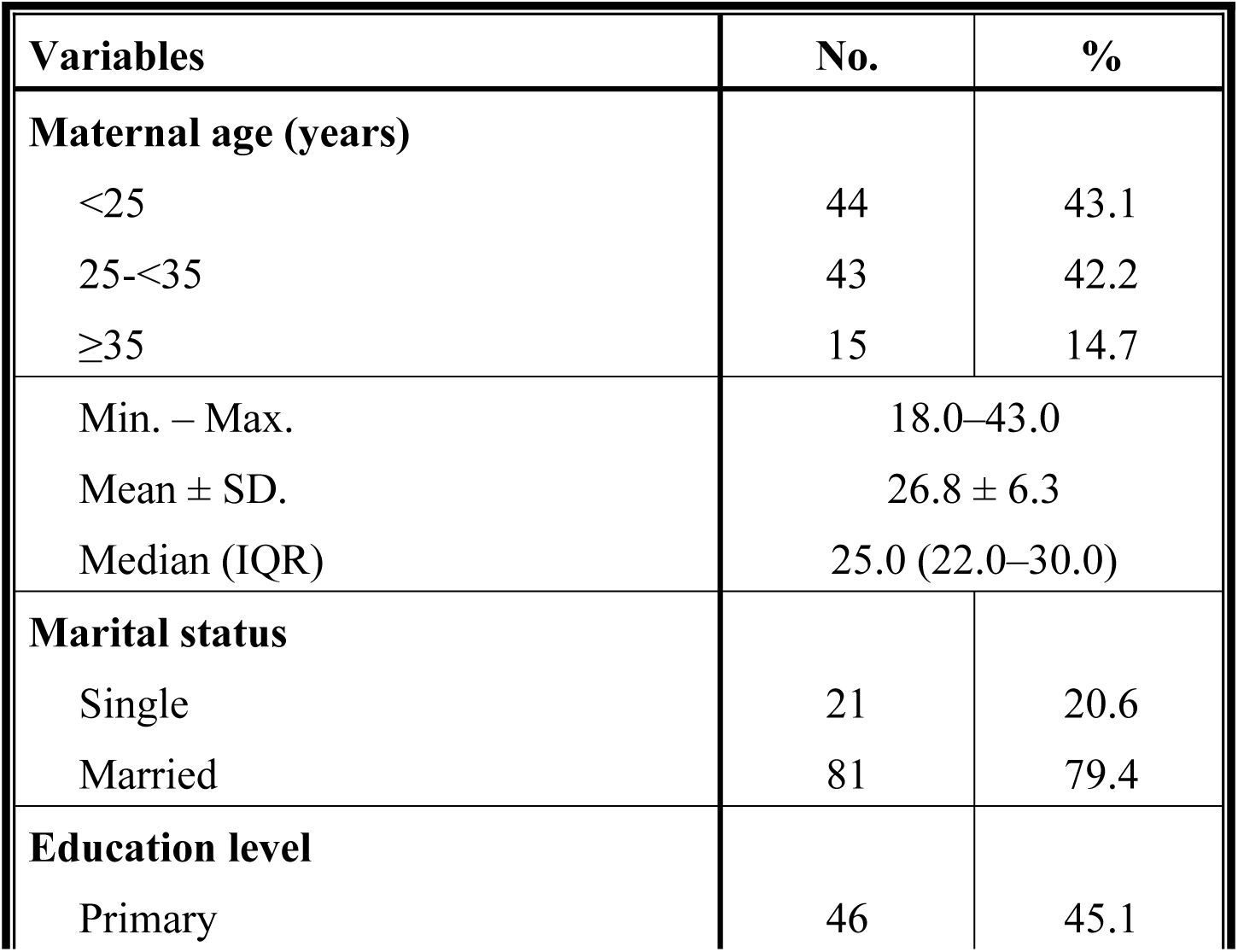
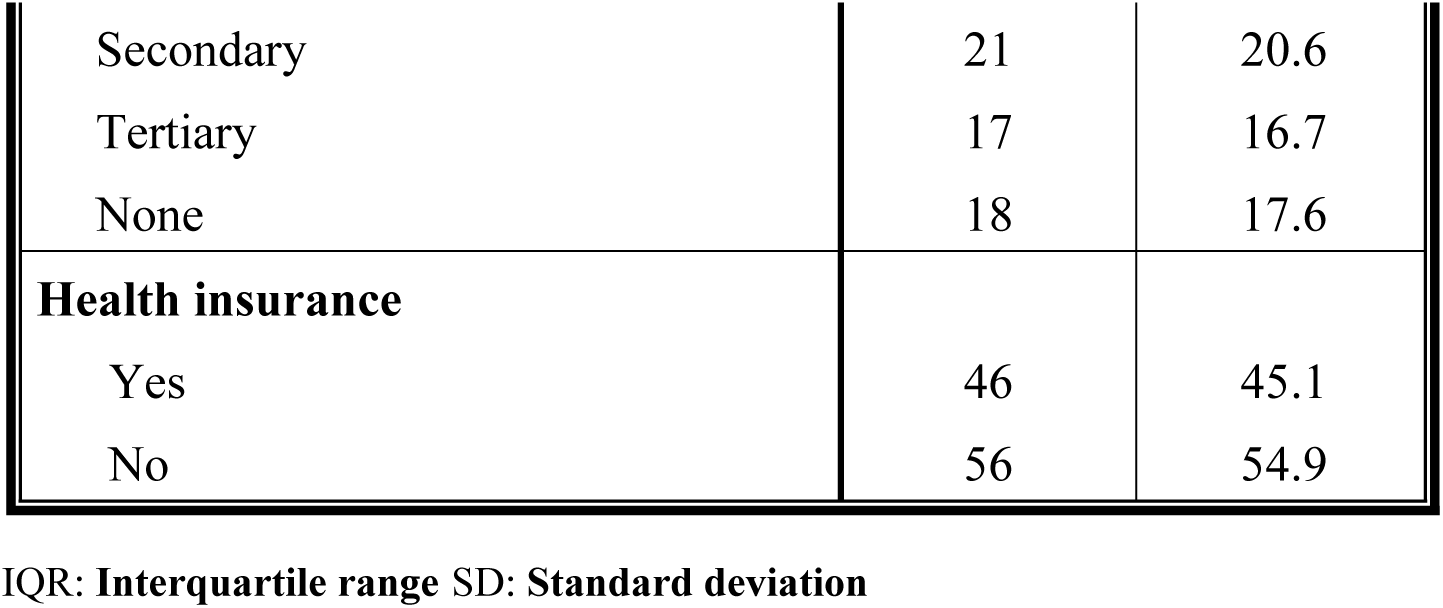
Maternal sociodemographic characteristics (n = 102)

| Variables | No. | % |
| --- | --- | --- |
| **Maternal age (years)** |  |  |
| <25 | 44 | 43.1 |
| 25-<35 | 43 | 42.2 |
| $\geq 35$ | 15 | 14.7 |
| Min. – Max. | 18.0–43.0 |  |
| Mean ± SD. | $26.8 \pm 6.3$ | |
| Median (IQR) | 25.0 (22.0–30.0) |  |
| **Marital status** |  |  |
| Single | 21 | 20.6 |
| Married | 81 | 79.4 |
| **Education level** |  |  |
| Primary | 46 | 45.1 |
| Secondary | 21 | 20.6 |
| Tertiary | 17 | 16.7 |
| None | 18 | 17.6 |
| **Health insurance** |  |  |
| Yes | 46 | 45.1 |
| No | 56 | 54.9 |

IQR: Interquartile range SD: Standard deviation

In maternal clinical characteristics, 23.5% of the patients were rhesus-negative, while the majority were reported to be rhesus-positive (see Table 2). Patients with comorbidities accounted for 54.9% of the sample, and 29.4% had anemia. Women who had positive screening tests for syphilis accounted for 11.8% of the sample, while women who had positive screening tests for human immunodeficiency virus (HIV) accounted for 9.8%.

**Table 2.** Maternal characteristics of interest.

| Maternal characteristics of interest | No. | % |
| --- | --- | --- |
| **Blood group** |  |  |
| A | 38 | 37.3 |
| B | 21 | 20.6 |
| AB | 3 | 2.9 |
| O | 40 | 39.2 |
| **Rhesus negative** |  |  |
| Yes | 24 | 23.5 |
| No | 78 | 76.5 |
| **Parity** |  |  |
| Primiparous | 41 | 40.2 |
| Multiparous ($\leq 4$ deliveries after 28 weeks GA) | 53 | 52.0 |
| Grand multiparous ($\geq 5$ deliveries after 28 weeks GA) | 8 | 7.8 |
| **History of pregnancy loss before 28 weeks GA** |  |  |
| Yes | 18 | 17.6 |
| No | 80 | 78.4 |
| NA | 4 | 3.9 |
| **Previous fetal death** |  |  |
| Yes | 24 | 23.5 |
| No | 78 | 76.5 |
| **Prior neonatal death** |  |  |
| Yes | 11 | 10.8 |
| No | 91 | 89.2 |
| **Number of fetuses** |  |  |
| Singleton | 83 | 81.4 |
| Twins (twin-to-twin transfusion syndrome) | 19 | 18.6 |
| **Maternal comorbidities** |  |  |
| Yes | 56 | 54.9 |
| No | 46 | 45.1 |
| **If yes** |  |  |
| Diabetes mellitus | 4 | 3.9 |
| Gestational diabetes | 6 | 5.9 |
| Sickle cell disease | 3 | 2.9 |
| Cardiac disease | 1 | 1.0 |
| Anemia | 30 | 29.4 |
| Chronic HTN | 12 | 11.8 |
| NA | 0 | 0.0 |
| **Antibody screening, for syphilis-positive individuals** |  |  |
| Yes | 12 | 11.8 |
| No | 90 | 88.2 |
| **Positive HIV screening** |  |  |
| Yes | 10 | 9.8 |
| No | 92 | 90.2 |
| **Mirror syndrome** |  |  |
| Yes | 16 | 15.7 |
| No | 86 | 84.3 |
| **Mode of delivery** |  |  |
| -Vaginal | 51 | 50.0% |
| -Cesarean section | 51 | 50.0% |

The majority of infants with hydrops fetalis (66.7%) had NIHF. The mean gestational age (standard deviation) was 34.1±3.9 weeks. Most newborns with hydrops fetalis (80.4%) had an Apgar score ≤ 7 at 5 minutes, and 35.3% of these babies had birth weights between 1600 grams and 2400 grams, followed by those with birth weights ≥3400 grams (30.4%). Seventy-three percent (73.5%) of the newborns with hydrops fetalis were admitted to the newborn unit (NBU), and 44.1% of them had a confirmed diagnosis of anemia (see Table 3). Only 42.2% of the newborns survived the perinatal period (Table 3).

**Table 3.**
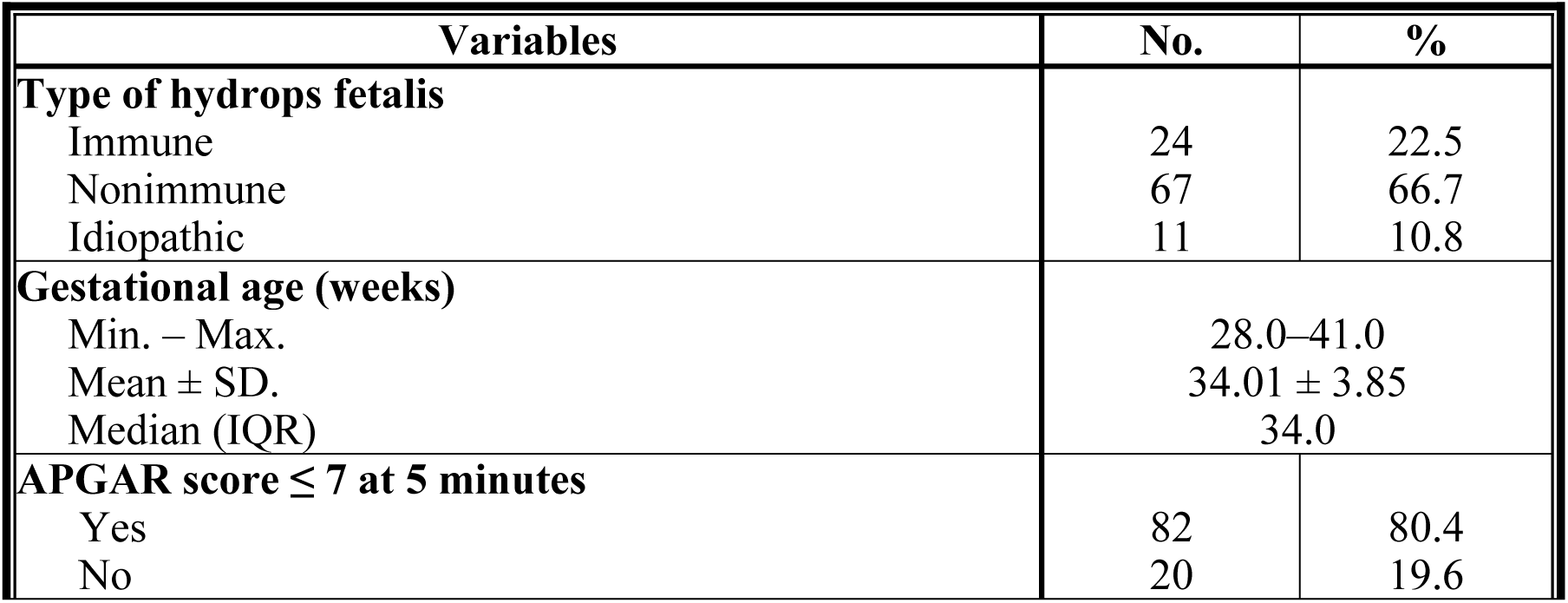
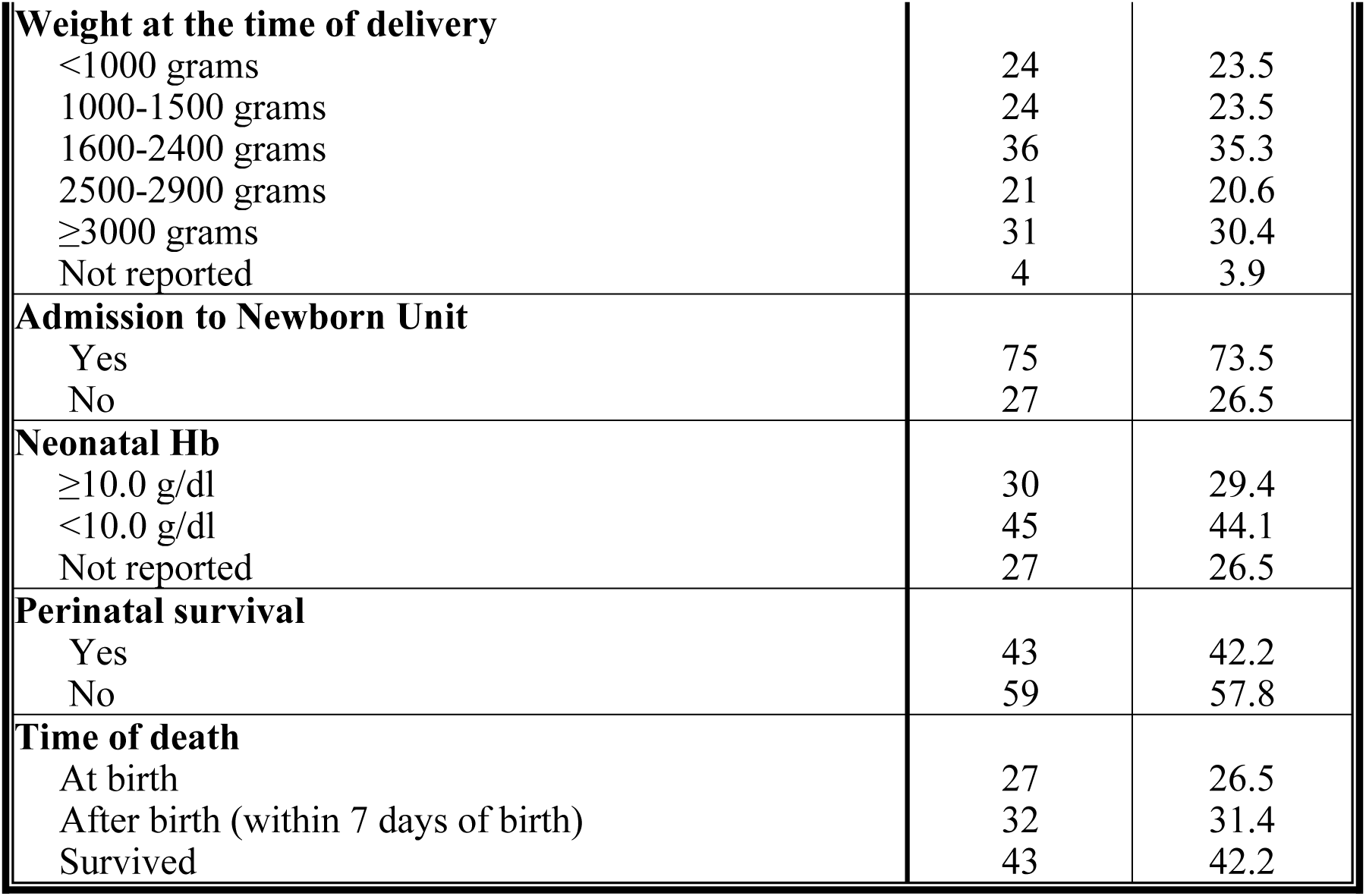
Perinatal outcome of interest.

Table 4 essentially shows the associations among survival, mode of delivery, and hydrops fetalis classification. The table shows that 67.4% of the surviving infants had NIHF, while 25.6% and 7.0% had IHF and idiopathic HF, respectively. Fifty-one percent (51.2%) vs. 48.8% of the surviving infants were born via the vaginal route. However, there was no statistically significant association between the mode of delivery and survival (pv=0.84). The infants with class II hydrops fetalis had a 47.5% rate of survival, whereas the rates were equally low for those with class III and IV hydrops fetalis. There was a statistically significant association between perinatal survival and the classification of hydrops fetalis (pv<0.001).

**Table 4.** Associations between survival and hydrops fetalis type, mode of delivery, and hydrops classification.

| Variables | Survival | | | | $\chi^2$ | P |
| --- | --- | --- | --- | --- | --- | --- |
|  | Yes<br>(n =43) |  | No<br>(n =59) |  |  |  |
|  | No. | % | No. | % |  |  |
| Type of hydrops fetalis |  |  |  |  |  |  |
| Immune | 12 | 25.6 | 12 | 20.3 | 1.608 | 0.447 |
| Nonimmune | 28 | 67.4 | 39 | 66.1 |  |  |
| Idiopathic | 3 | 7.0 | 8 | 13.6 |  |  |
| **Mode of delivery** |  |  |  |  |  |  |
| Vaginal | 22 | 51.2 | 29 | 49.2 | 0.040 | 0.841 |
| Cesarean | 21 | 48.8 | 30 | 50.8 |  |  |
| **Class of the patients** |  |  |  |  |  |  |
| Class I: mild hydrops fetalis | 3 | 7.5 | 1 | 1.8 | 42.264* | <sup>MC</sup>p<br><0.001* |
| Class II: moderate hydrops fetalis | 19 | 47.5 | 2 | 3.5 |  |  |
| Class III: moderately severe hydrops fetalis | 9 | 22.5 | 6 | 10.5 |  |  |
| Class IV: severe hydrops fetalis | 9 | 22.5 | 48 | 84.2 |  |  |

$\chi^2$ : Chi-square test
p: p value for comparison between the studied categories
\*: Statistically significant at $p \leq 0.05$

The absence of congenital malformation was significantly associated with survival (76.7% vs. 23.3%, pv=008). In addition, normal Doppler findings were found to be significantly associated with perinatal survival (pv=0.001), as shown in Table 5.

**Table 5.** Associations between survival and ultrasound findings (n = 102)

| Variables | Survival | | | | $\chi^2$ | p |
| --- | --- | --- | --- | --- | --- | --- |
|  | Yes<br>(n =43) |  | No<br>(n =59) |  |  |  |
|  | No. | % | No. | % |  |  |
| **Presence of congenital abnormalities** |  |  |  |  |  |  |
| Yes | 10 | 23.3 | 29 | 49.2 | 7.063* | 0.008* |
| No | 33 | 76.7 | 30 | 50.8 |  |  |
| **If congenital abnormalities** |  |  |  |  |  |  |
| Congenital hydrothorax | 0 | 0.0 | 1 | 1.7 | 10.849 | <sup>MC</sup>p=<br>0.104 |
| Skeletal dysplasia | 1 | 2.3 | 8 | 13.6 |  |  |
| Urinary abnormalities | 1 | 2.3 | 0 | 0.0 |  |  |
| Gastrointestinal abnormalities | 2 | 4.7 | 5 | 8.5 |  |  |
| Fetal tumors | 3 | 7.0 | 4 | 6.8 |  |  |
| Twin-to-twin transfusion syndrome | 1 | 2.3 | 7 | 11.9 |  |  |
| No fetal anomaly | 17 | 39.5 | 16 | 27.1 |  |  |
| Not indicated | 18 | 41.9 | 18 | 30.5 |  |  |
| **Doppler study** |  |  |  |  |  |  |
| UA increased S/D ratio | 3 | 7.0 | 0 | 0.0 | 20.232* | <sup>MC</sup>p<br><0.001* |
| UA A/R EDF | 11 | 25.6 | 29 | 49.2 |  |  |
| MCA PI <10 percentile | 18 | 41.9 | 11 | 18.6 |  |  |
| MCA PSV>1.5 MoM | 6 | 14.0 | 4 | 6.8 |  |  |
| DV A/R a-wave | 3 | 7.0 | 15 | 25.4 |  |  |
| Not indicated | 2 | 4.7 | 0 | 0.0 |  |  |

$\chi^2$ : **Chi-square test** MC: **Monte Carlo** FE: **Fisher exact**
p: p value for comparison between the studied categories
\*: Statistically significant at $p \leq 0.05$

**Table 6.** Univariate and multivariate logistic regression analysis.

| Variables | Univariate |  | #Multivariate |  |
| --- | --- | --- | --- | --- |
|  | p | OR (LL – UL 95% CI) | p | OR (LL – UL 95% CI) |
| $\geq 3000$ grams | **0.02\*** | 0.35(0.14–0.85) | 0.02 | 0.35(0.14–0.87) |
| Admission to the NBU | **0.04\*** | 2.55(1.01–6.39) | 0.05 | 2.51(0.98–6.46) |

OR: odds ratio
CI: Confidence interval LL: Lower limit UL: Upper limit
#: All variables with $p < 0.05$ were included in the multivariate analysis.
\*: Statistically significant at $p \leq 0.05$

The presence of maternal comorbidities, especially anemia was significantly associated with reduced survival rates (pv=0.035). There was a significant association between mirror syndrome and reduced perinatal survival (pv=0.009) as shown in supplementary table 2.

### Univariate and multivariate logistic regression

Finally, the univariate analysis showed that a birth weight ≥3000 grams was associated with better perinatal outcomes (0.3%, pv=0.02, 95% CI=0.1-0.8). Even with multivariate logistic regression a birth weight ≥3000 grams improved the perinatal survival with a relative risk of 0.4 (pv=0.02, 95% CI=0.1-0.9). Similarly, admission to the NBU was associated with a 2.5 risk reduction in mortality (pv=0.04, 95% CI=1.0–6.4). After multivariate logistic regression, admission to the NBU reduced perinatal mortality by 2.5 (pv=0.05, 95% CI= 1.0–6.5). In this study, the 2 variables (birth weight and admission to the newborn care unit) appeared to determine the perinatal outcomes of hydrops fetalis infants.

## Comments

### Principal findings

The principal findings of this study are as follows: 1) Hydrops fetalis is associated with reduced perinatal survival, and several factors contribute to the observed outcomes, including the severity of the disease (class III and IV), low birth weight, type of hydrops fetalis, and maternal comorbidities. The observed outcomes were not statistically associated with the mode of delivery. 2) Mirror syndrome, which refers to a clinical entity that can occur in a pregnancy complicated by fetal hydrops, significantly reduces the survival rate of infants with hydrops fetalis. 3) Admission to the NBU or NICU for advanced neonatal care improves perinatal survival, as well as birth weight. It is reasonable to gain time of delivery for classes I and II at a critical gestational age (<30 weeks) with nearly normal UA Doppler or MCA PI. 4) Cesarean delivery does not improve the perinatal outcomes of infants with hydrops fetalis; however, vaginal delivery seems to be beneficial to mothers and fetuses. 5) High survival rates are observed among infants with PHIOMoD class I and II hydrops fetalis. However, the number of participants in class I and II was too small compared to those in classes III and IV. Therefore, further study with an equal number of participants is recommended to validate the PHIOMoD predictive tool.

### Results in the context of what is known

There is evidence that hydrops fetalis confers a high risk of morbidity and mortality^8,10^. Our study, which substantially extends the existing knowledge in the field, revealed that hydrops fetalis is associated with poor survival rates. There is also evidence that NIHF is the most prevalent type, and its etiology is multifactorial^11^.

There is evidence that the prognosis depends on the underlying cause of the syndrome^8^. However, it should be acknowledged that the severity of the disease, in-utero management status, maternal comorbidities, birth weight, Apgar score, newborn anemia status, and management after birth are all major determinants of survival rather than the underlying cause alone^12,13^. Mirror syndrome, which refers to a maternal clinical characteristic, is reported to be present in some cases^14,16^. The incidence of mirror syndrome is unclear and may not be reported or may be wrongly attributed to severe preeclampsia. Although there was no maternal death from mirror syndrome reported in this study, the literature shows a maternal mortality rate of approximately 20% secondary to pulmonary edema^17,18^. With treatment for fetal hydrops, maternal symptoms have been noted to resolve^19,20^.

Recently, evidence has also shown that most women with pregnancies complicated by hydrops fetalis delivery by cesarean section^8^. However, this study indicates that the mode of delivery does not improve perinatal outcomes for these infants⁸.

### Clinical implications

These findings showed that cesarean delivery does not improve the perinatal outcomes of infants with hydrops fetalis in low-resource settings country. Although there was increased survival in PHIOMoD class I and II patients and reduced survival in class III and IV patients, vaginal delivery remains the preferred mode of delivery for pregnancies complicated by hydrops fetalis. Cesarean section should be performed for obstetric indications, either for maternal or fetal concerns (if class I or II disease). There is no optimal mode of delivery for pregnancies complicated by hydrops fetalis and congenital malformation. However, options for management during the prenatal, perinatal, intrapartum, and neonatal periods need to be thoroughly discussed so that families can make informed decisions^21^. A multidisciplinary approach is usually needed once a decision has been made to optimize fetal outcomes and to plan for the timing and location as well as the mode of delivery. Vaginal delivery is preferred, and cesarean section should be considered for maternal concerns, including protracted labor, abruptio placentae with maternal hemodynamic instability, previous cesarean section for a recurrent indication or ≥2 scars, uncontrolled eclamptic seizures, complete placenta previa, and malpresentation^²²^. In low-resource settings, morally, it is not acceptable to perform a cesarean section where there is a very high probability of a severe fetal anomaly being diagnosed and a very high probability of neonatal death after delivery. The PHIOMoD Index remains an important tool for assisting obstetricians in decision-making regarding the mode of delivery for women with pregnancies complicated by hydrops fetalis.

### Research implications

Data regarding the perinatal outcomes of infants with hydrops fetalis and the PHIMoD classification are needed to extend the knowledge in the field. Although the etiology and pathophysiology of the condition have been classified, little is known about the impact of the mode of delivery on improving infant survival without leading to unnecessary sequelae in mothers. In addition, prospective observational data combined with in-utero management and perinatal outcomes are needed to assess the performance of the PHIOMoD classification, especially for hydrops fetalis classes III and IV. The goal is to improve the counseling of affected women and reduce the cesarean section rates indicated for fetal concerns, especially hydrops fetalis.

### Strengths and limitations

The main strength of this study is the use of a cohort of women with pregnancies complicated with hydrops fetalis, in which the diagnostic criteria were defined. A multidisciplinary team used scientific evidence to create a comprehensive classification of hydrops fetalis based on severity and predict outcomes by mode of delivery based on the classification. The standard of evidence for the optimal management of pregnant women whose fetuses are diagnosed with hydrops fetalis and management outcomes were also included. The variability in practice between the various hospitals worldwide, experience, differences in resources, and access to intrauterine management and neonatal care were acknowledged.

The limitations of this study included the following: 1) Ultrasound findings were reported by different sonographers and radiologists within the institution who do not probably have the same experience. This could result in bias and inaccurate findings to correctly allocate patients. 2) The participants were not matched, especially for gestational age, to reduce bias in perinatal outcomes. 3) The PHIMoD study tool has not been externally validated, and there is a need for validation by independent researchers to determine the reproducibility and generalizability of the prediction model for new and different patients. 4) the study was done in low low-resource setting context and the PHIOMoD index is useful to improve care through decision-making in low-resource settings only where there is limited access to intrauterine fetal therapy. 5) genetic testing, karyotyping, and most serological tests were not performed for the completeness of investigations due to financial constraints and limited resources. This implied defining idiopathic hydrops fetalis. 6) intrauterine intervention or transfusion was not performed on those who could ideally benefit due to limited resources. 7) this was a retrospective study, with questions related to missing information and incompleteness of key data that may not be answered. However, the PHIOMoD II prospective cohort study will help to mitigate the limitations.

## Conclusion

Cesarean section delivery does not improve the perinatal outcomes of newborns with severe hydrops fetalis who do not receive intra-uterine intervention or transfusion. It should be considered only for maternal concerns. Birth weight and advanced newborn care at the newborn unit or newborn intensive care unit improve perinatal outcomes. The PHIOMoD tool may be a good predictor of perinatal outcomes based on the mode of delivery, especially in low-resource settings.

## Data Availability

N/A

## Acknowledgment

The authors are sincerely grateful to Isaac Mwanza, the record office, and the healthcare providers of the Moi Teaching and Referral Hospital, whose support and contributions have been instrumental in completing this study.

## Funding

This study was funded by the Moi Teaching and Referral Hospital Intramural Research Fund (MIRF).

## Conflicts of interest

The authors report no conflicts of interest.

## Key findings

- The overall perinatal survival by mode of delivery was similar for infants delivered vaginally and by cesarean section, accounting for 51.2% and 48.8%, respectively.
- A birth weight ≥3000 grams was associated with increased survival (0.3%, pv=0.02, 95% CI=0.14-0.85). Admission to the newborn unit was associated with a risk relative of 2.5 increase in survival (pv=0.04, 95% CI=1.0–6.4).

## What does this add to what is known?

This study provides evidence that hydrops fetalis is associated with reduced survival regardless of the mode of delivery. The clinical classification used in this study, which is based on ultrasonographic findings of fetal anomalies, helps assist in decision-making regarding the mode of delivery for infants with hydrops fetalis.

## SUPPLEMENTARY TABLES FOR PHIOMod STUDY

**Table 1.**
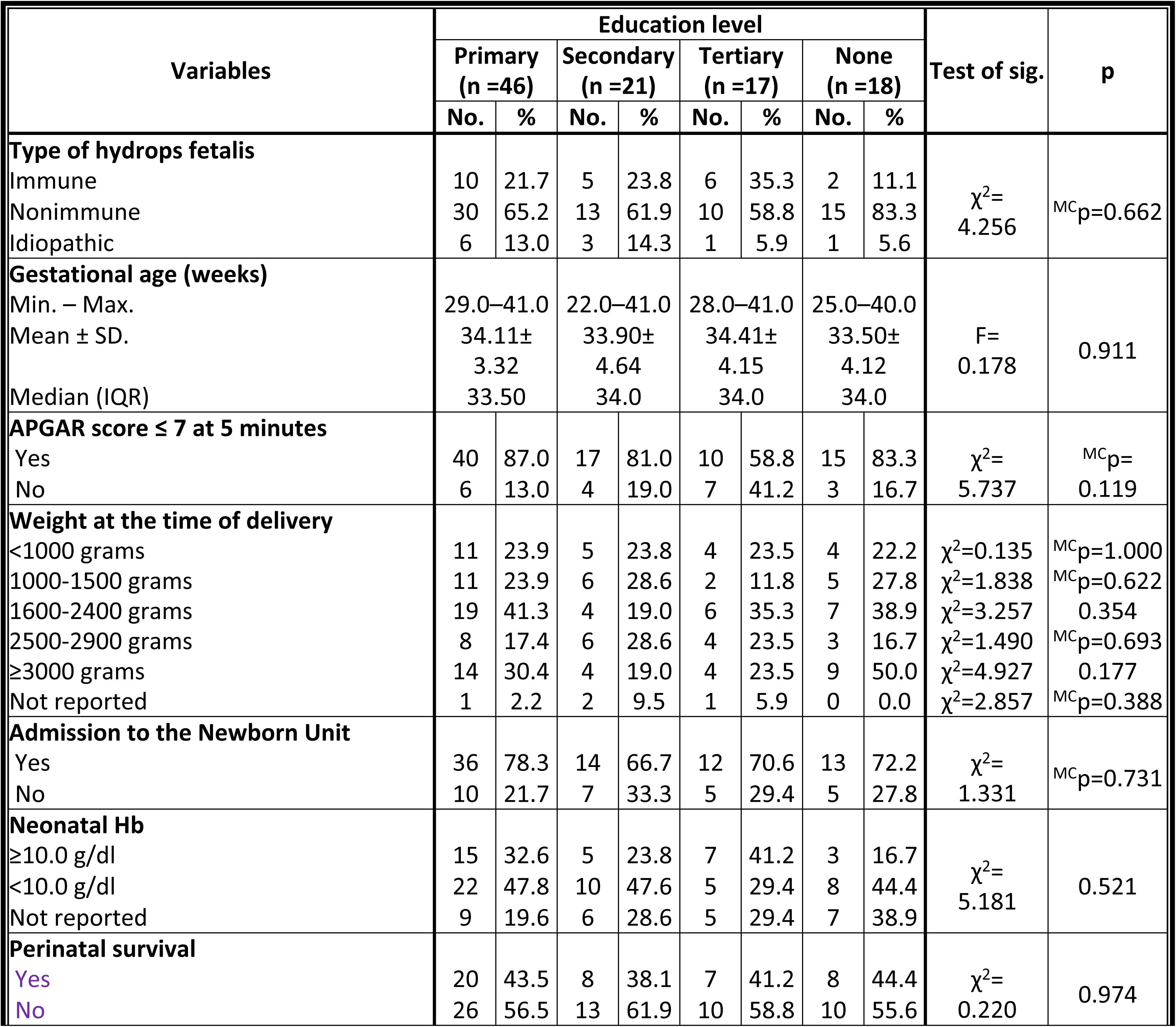
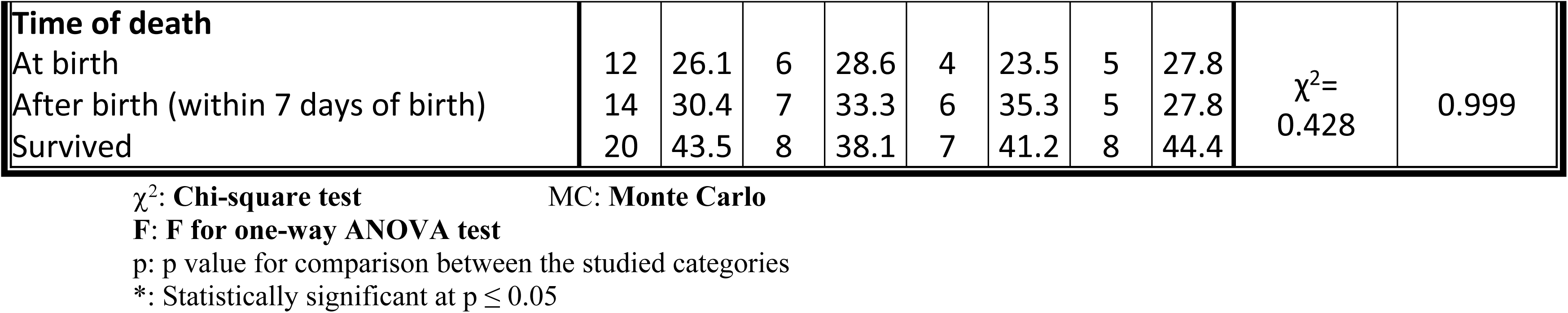
Associations between maternal education level and perinatal outcomes of interest (n =102)

**Table 2.** Associations between survival and maternal clinical characteristics (n = 102)

| Maternal characteristics of interest | Survival | | | | $\chi^2$ | p |
| --- | --- | --- | --- | --- | --- | --- |
|  | Yes<br>(n =43) |  | No<br>(n =59) |  |  |  |
|  | No. | % | No. | % |  |  |
| Blood group |  |  |  |  | 6.515 | <sup>MC</sup>p=0.072 |
| A | 13 | 30.2 | 25 | 42.4 |  |  |
| B | 14 | 32.6 | 7 | 11.9 |  |  |
| AB | 1 | 2.3 | 2 | 3.4 |  |  |
| O | 15 | 34.9 | 25 | 42.4 |  |  |
| Rhesus negative |  |  |  |  | 0.792 | 0.374 |
| Yes | 12 | 27.9 | 12 | 20.3 |  |  |
| No | 31 | 72.1 | 47 | 79.7 |  |  |
| Parity |  |  |  |  | 2.516 | <sup>MC</sup>p=0.288 |
| Primiparous | 19 | 44.2 | 22 | 37.3 |  |  |
| Multiparous | 19 | 44.2 | 34 | 57.6 |  |  |
| Grand multiparous | 5 | 11.6 | 3 | 5.1 |  |  |
| History of pregnancy loss before 28 weeks GA |  |  |  |  | 2.552 | <sup>MC</sup>p=0.285 |
| Yes | 9 | 20.9 | 9 | 15.3 |  |  |
| No | 31 | 72.1 | 49 | 83.1 |  |  |
| NA | 3 | 7.0 | 1 | 1.7 |  |  |
| Previous fetal death |  |  |  |  | 0.279 | 0.597 |
| Yes | 9 | 20.9 | 15 | 25.4 |  |  |
| No | 34 | 79.1 | 44 | 74.6 |  |  |
| Prior neonatal death |  |  |  |  | 0.055 | <sup>FE</sup>p=1.000 |
| Yes | 5 | 11.6 | 6 | 10.2 |  |  |
| No | 38 | 88.4 | 53 | 89.8 |  |  |
| Number of fetuses |  |  |  |  | 2.372 | 0.124 |
| Singleton | 32 | 74.4 | 51 | 86.4 |  |  |
| Twins | 11 | 25.6 | 8 | 13.6 |  |  |
| Maternal comorbidity |  |  |  |  | 11.799* | <sup>MC</sup>p=0.035* |
| Diabetes mellitus | 1 | 2.3 | 3 | 5.1 |  |  |
| Gestational diabetes | 6 | 14.0 | 0 | 0.0 |  |  |
| Sickle cell disease | 1 | 2.3 | 2 | 3.4 |  |  |
| Cardiac disease | 1 | 2.3 | 0 | 0.0 |  |  |
| Anemia | 11 | 25.6 | 19 | 32.2 |  |  |
| Chronic HTN | 3 | 7.0 | 9 | 15.3 |  |  |
| Antibody screening, for syphilis-positive individuals |  |  |  |  | 0.434 | 0.510 |
| Yes | 4 | 9.3 | 8 | 13.6 |  |  |
| No | 39 | 90.7 | 51 | 86.4 |  |  |
| Positive HIV screening |  |  |  |  |  |  |
| Yes | 3 | 7.0 | 7 | 11.9 | 0.672 | <sup>MC</sup>p=0.513 |
| No | 40 | 93.0 | 52 | 88.1 |  |  |
| **Mirror syndrome** |  |  |  |  |  |  |
| Yes | 2 | 4.7 | 14 | 23.7 | 6.845* | 0.009* |
| No | 41 | 95.3 | 45 | 76.3 |  |  |

$\chi^2$ : Chi-square test MC: Monte Carlo FE: Fisher exact
p: p value for comparison between the studied categories

**Figure 1.**
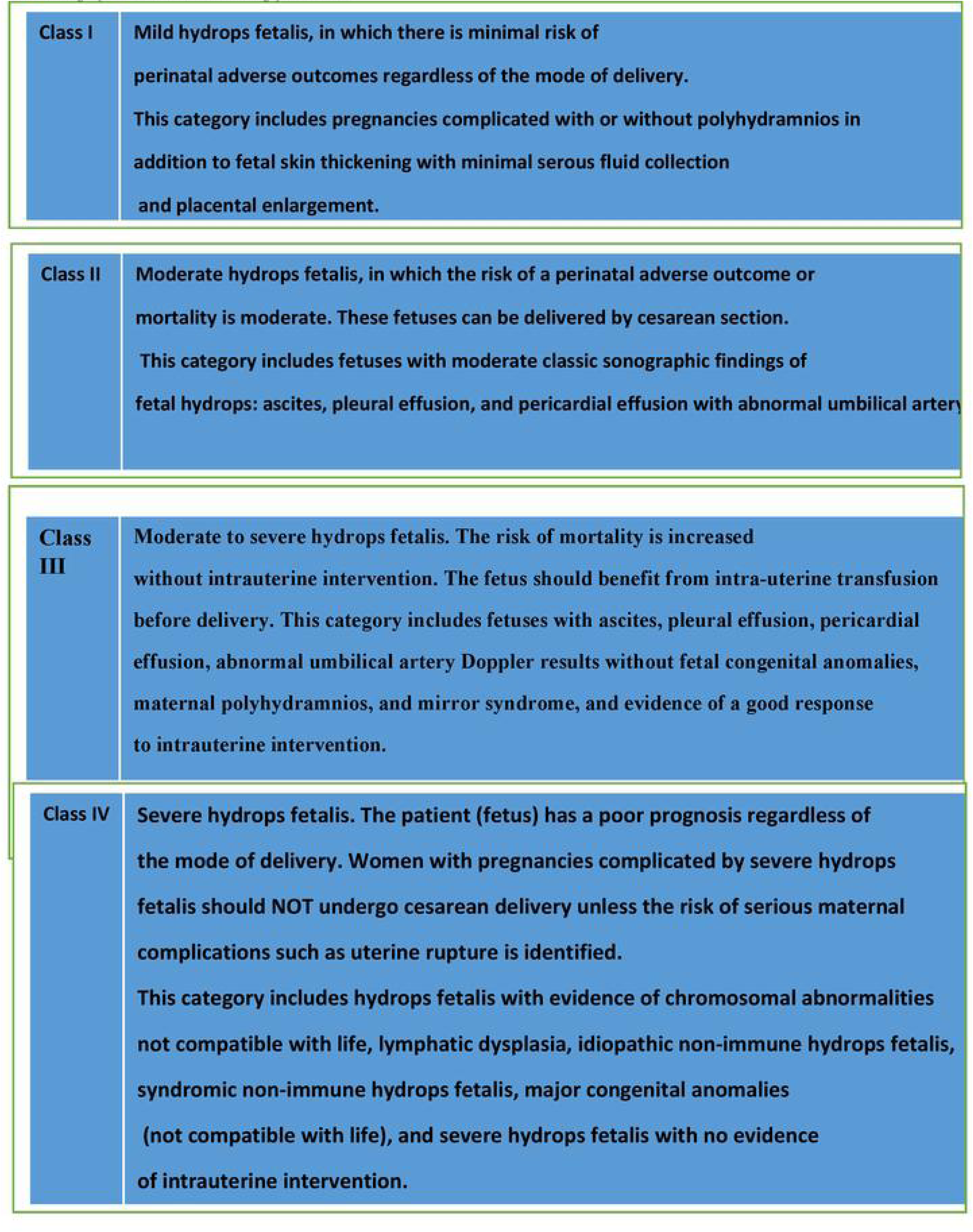
Prediction of perinatal outcomes of infants with hydrops fetalis by mode of deliverv (PHIOMoD Studv) index.

**Figure 2.**
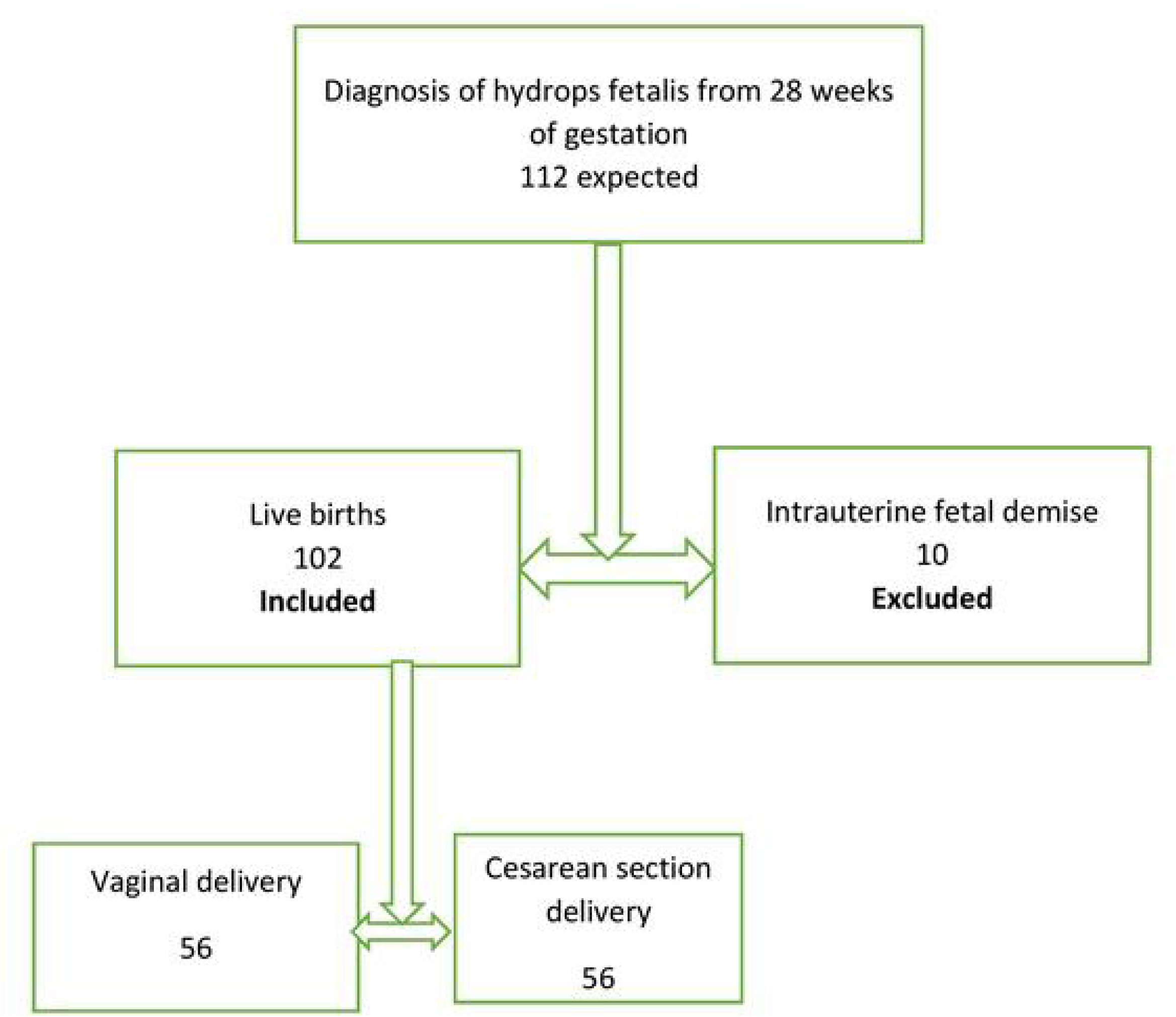
Flowchart of participants’ recuirtment

## Notes

### Competing Interest Statement

The authors have declared no competing interest.

### Funding Statement

Yes

### Author Declarations

This study was approved by the Moi Teaching and Referral Hospital- Moi University Institutional Research and Ethics Committee (IREC) under the number FAN 0003998

